# Prevalence of SARS-CoV-2 antibodies among workers of the public higher education institutions of Porto, Portugal

**DOI:** 10.1101/2021.02.28.21252628

**Authors:** Paula Meireles, Joana Amaro, Joana Pinto Costa, Mariana Mendes Lopes, Tatiana Varandas, Pedro Norton, João Tiago Guimarães, Milton Severo, Henrique Barros

**Affiliations:** EPIUnit–Instituto de Saúde Pública, Universidade do Porto, Rua das Taipas, nº 135, 4050-600 Porto, Portugal; Serviço de Saúde Ocupacional, Centro Hospitalar Universitário de São João, Porto, Portugal; Serviço de Patologia Clínica, Centro Hospitalar Universitário de São João, Porto, Portugal; Faculdade de Medicina, Universidade do Porto, Alameda Prof. Hernâni Monteiro, 4200-319 Porto, Portugal

## Abstract

**Objectives:** To assess the prevalence of SARS-CoV-2 specific immunoglobulin (Ig) M and IgG antibodies among workers of the three public higher education institutions of Porto, Portugal, up to July 2020.

**Methods:** A rapid point of care test for specific IgM and IgG antibodies of SARS-CoV-2 was offered to all workers. Testing was performed to and a questionnaire was completed by 4592 workers on a voluntary basis. We computed the apparent IgM, IgG, and combined IgM or IgG prevalence, along with the true prevalence and 95% credible intervals (95% CI) using Bayesian inference.

**Results:** We found an apparent prevalence of 3.1% for IgM, 1.0% for IgG, and 3.9% for either antibody class. The estimated true prevalence was 2.0% (95% CI 0.1-4.3) for IgM, 0.6% (95% CI 0.0-1.3) for IgG and 2.5% (95% CI 0.1-5.3) for IgM or IgG. A SARS-CoV-2 molecular diagnosis was reported by 21 (0.5%) workers, and of these, 90.5% had a reactive IgG result. Seroprevalence was higher among those reporting known contacts with confirmed cases, having been quarantined, having a previous molecular negative test, or having had symptoms.

**Conclusions:** The seroprevalence among workers from the three public higher education institutions of Porto after the first wave of the SARS-CoV-2 infection was relatively low. However, the estimated true seroprevalence was approximately five times higher than the reported SARS-CoV-2 infection based on a molecular test result.

## INTRODUCTION

The SARS-CoV-2 infection can cause very severe disease, particularly among individuals with underlying conditions. Commonly it progresses unnoticed with few or no symptoms [1] – additional limited testing capacity has led to a variable and mostly unknown undiagnosed rate.

Seroprevalence studies are based on the identification of SARS-CoV-2 specific antibodies. In this case of an emergent agent, the entire population is initially susceptible. Therefore, the presence of specific antibodies provides estimates of the cumulative incidence of infection. In SARS-CoV-2 infection, almost all of the infected individuals seroconvert within 2-3 weeks.[2-4]

Diseases with an impact on the working population cause very high individual and societal costs. Activities where inter-personal contact is inevitable, structural or individual lack of compliance with preventive measures, sharing the same office or canteen space, and meeting in overcrowded rooms, may increase the SARS-CoV-2 infection in the workplace.[5] Preventive measures include the use of face masks, hand sanitizers, increased distance between workers, scattered working hours, or working from home. The latter has been deemed mandatory in Portugal from March 18 to June 30, 2020. The return to workplace activities provided an excellent opportunity to obtain data on serum status. Therefore, we aimed to assess the prevalence of SARS-CoV-2 specific immunoglobulin (Ig) M and IgG antibodies among workers of public higher education institutions of Porto, Portugal, from May to July 2020.

## METHODS

All workers of the three public higher education institutions of Porto were offered a serological point-of-care test for SARS-CoV-2 specific IgM and IgG antibodies, from May 21 to July 31, 2020. Participation was voluntary, and scheduling was initiated by the workers. At the day of testing workers were invited to answer to two questionnaires – one to evaluate clinical aspects, conducted by the trained researcher who performed the test, and another self-administered to address sociodemographic characteristics.

The clinical questionnaire included information on comorbidities, contacts with confirmed SARS-CoV-2 cases in the previous two weeks, symptoms since the beginning of 2020 (categorized into asymptomatic; moderately symptomatic: one or two of the following symptoms cough, dyspnea, odynophagia, headache, vomiting or nausea, diarrhea, asthenia, or fever; and symptomatic: at least three of the listed symptoms, or dysgeusia or anosmia), and previous SARS-CoV-2 diagnostic tests. The self-administered questionnaire inquired about gender identity, nationality, educational level, occupation, currently working from home, self-perception of having been infected, travelling abroad since December 2019, contacts with confirmed SARS-CoV-2 cases and having been quarantined since January 2020

Participants provided written informed consent to all procedures. The study protocol was approved by the ethics committee of the Institute of Public Health of the University of Porto (ID 20154).

### SARS-CoV-2 specific IgM and IgG antibodies determination and follow-up

Two point-of-care tests were used – the STANDARD Q COVID-19 IgM/IgG Duo used from May 21 to July 10, n=3987 (manufacturer reported sensitivity of 92.6% eight days after symptom onset and specificity of 96.5% for both IgG and IgM); and the STANDARD Q COVID-19 IgM/IgG Combo from July 10 to July 31, n=605 (manufacturer reported sensitivity of 94.5% seven or more days after symptom onset and specificity of 95.7% for both IgG and IgM).

Participants reporting current symptoms or high-risk contact in the previous 14 days (spending more than 15 minutes within two meters of a confirmed case without any personal protective equipment), and those with a reactive result only for IgM were offered a referral to a reverse transcriptase-polymerase chain reaction (RT-PCR) test, scheduled within one working day.

### Participants

148 workers were tested from the Nursing School of Porto (ESEP), 816 from the Polytechnic of Porto (P.Porto) and 3628 from the University of Porto (U.Porto). Participants’ characteristics are presented in Table 1.

**Table 1:**
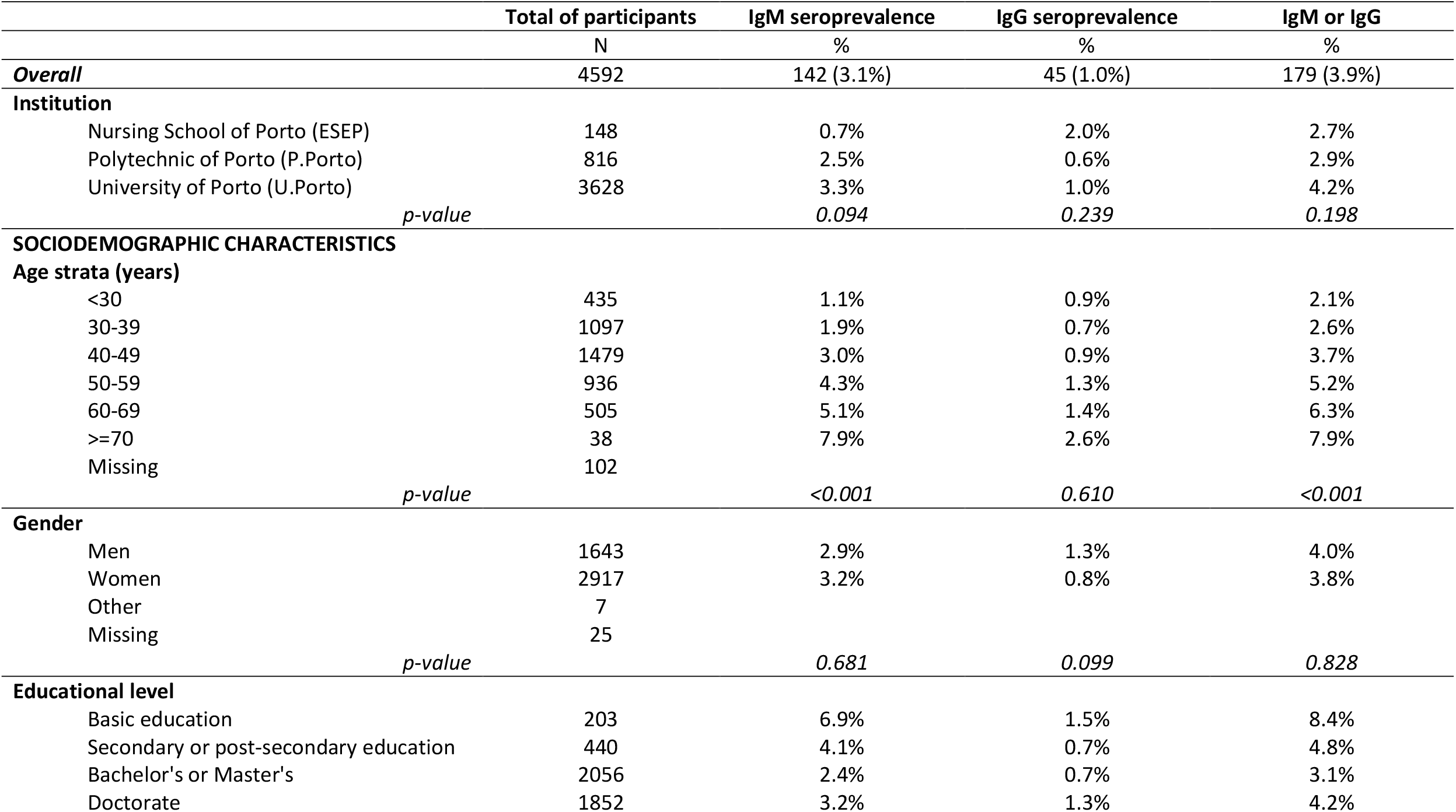
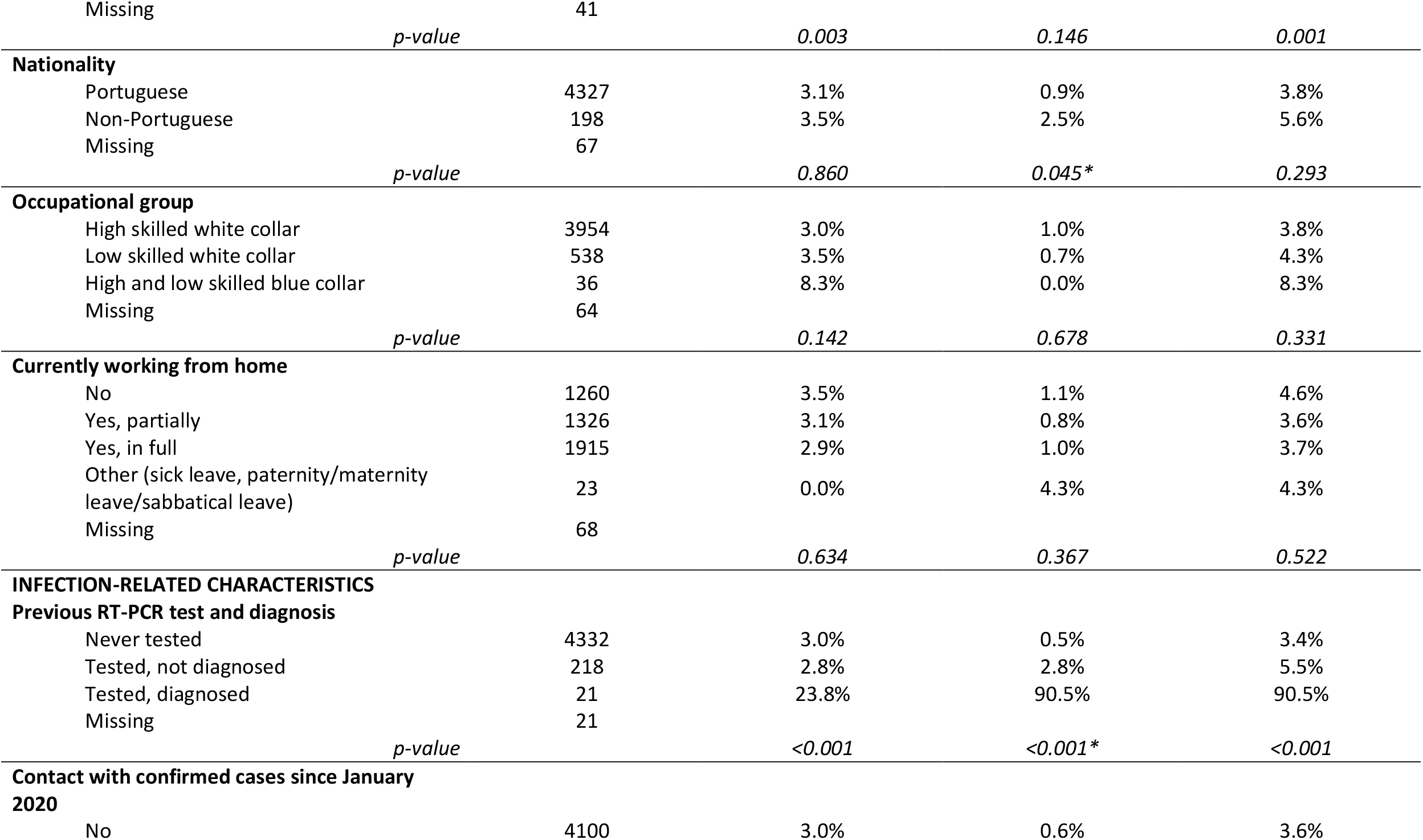
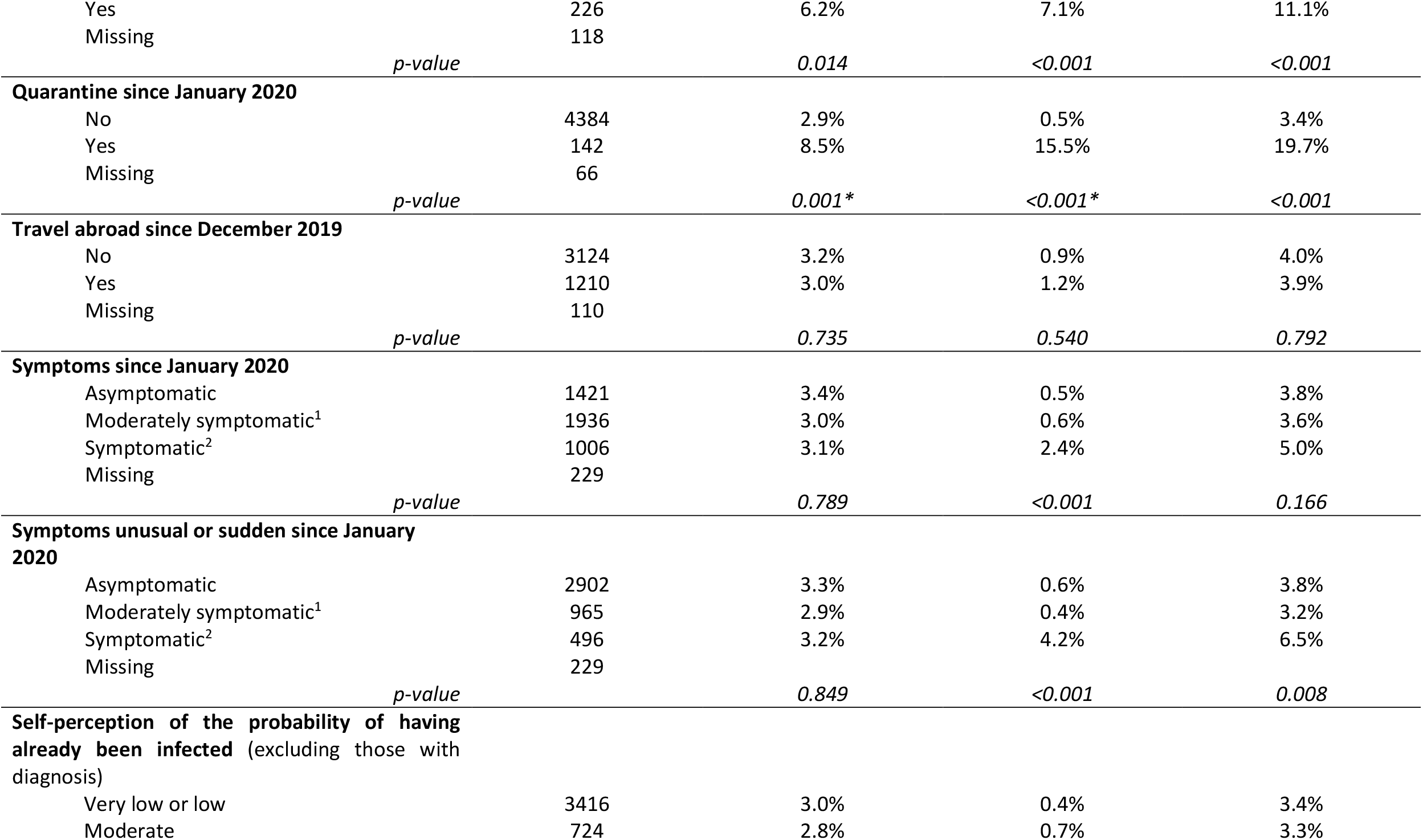
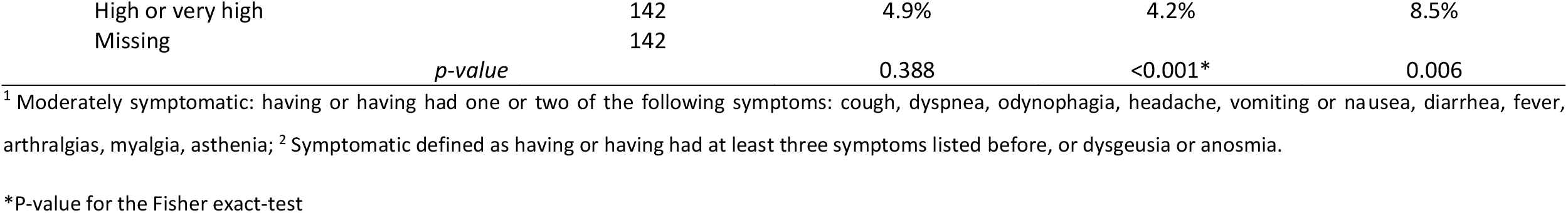
Characteristics of participants, IgM, IgG and IgM or IgG apparent seroprevalence according to those characteristics among workers from the public higher education institutions of Porto, Portugal, assessed from May to July 2020

### Statistical analysis

The apparent seroprevalence was computed as the proportion of individuals with a reactive result in the IgM or IgG band. We compared groups using the Pearson Chi-Square or the Fisher-exact test, when the assumptions for the chi-square test did not hold. P-values lower than 0.05 were considered statistically significant.

We estimated the true prevalence and 95% credible intervals (95% CI) using Bayesian inference, considering a uniform prior distribution for sensitivity ranging from 0.65 to 0.97, and specificity between 0.83 and 1. Estimates were obtained using the ‘rjags’ package in R.

## RESULTS

We tested 4592 workers; 142 (3.1%) were reactive for IgM, 45 (1.0%) for IgG, and 179 (3.9%) for at least one. The estimated true prevalence was 2.0% (95 %CI 0.1-4.3%) for IgM, 0.6% (95% CI 0.0-1.3%) for IgG and 2.5% (95% CI 0.1-5.3%) for IgM or IgG.

Table 1 presents the IgM, IgG, and IgM or IgG apparent seroprevalence according to the characteristics of the workers of all public higher education institutions of Porto. IgM seroprevalence increased significantly with age, and it was higher among those with the lowest educational levels. No gender or nationality differences were found, as well as in working from home status. IgM prevalence was higher among those with a previous diagnosis of SARS-CoV-2 infection (23.8%) when compared with those never tested (3.0%) or those who tested negative (2.8%). IgM was also higher among those with previous contact with confirmed cases (6.2% vs. 3.0%) and in those quarantined (8.5% *vs*. 2.9%). Travelling abroad, symptoms perceived as unusual or sudden, and self-perception of having been infected were not associated with IgM presence.

IgG seroprevalence did not differ according to age, gender, educational level, occupational group, or working from home. Non-Portuguese workers had a higher IgG seroprevalence (2.5% *vs*. 0.9%). Almost all of those diagnosed with SARS-CoV-2 infection had a reactive IgG test (19/21; 90.5%), and IgG prevalence was also higher among those who tested negative (2.8%) than those never tested (0.5%). IgG seroprevalence was higher among those with known contact with a confirmed case (7.1% *vs*. 0.6%), and among those who had been quarantined (15.5% *vs*. 0.5%). Those who reported any symptom had a higher IgG seroprevalence, particularly those symptomatic and with unusual or sudden onset of symptoms (4.2% *vs*. 0.4% among moderately symptomatic *vs*. 0.6% among symptomatic). IgG reactivity was more common in participants perceived the probability of having already been infected as high or very high.

A referral to a RT-PCR test was offered to 145 participants. Four refused (all IgM reactive). Of the remaining 141, 130 were IgM reactive and IgG non-reactive (15 also presented symptoms), three were IgM reactive and IgG reactive, seven presented symptoms (IgM and IgG non-reactive), and one had high-risk contacts (IgM and IgG non-reactive). Of the 141 RT-PCR tests, one was positive and corresponded to a worker referred due to symptoms and non-reactive results for IgM and IgG. All 133 workers with a reactive result only for IgM had a negative RT-PCR test.

## DISCUSSION

We found a 3.9% seroprevalence of IgM and/or IgG among workers of the three public higher education institutions of Porto, and a true prevalence of 2.5% (95% CI 0.1-5.3). The apparent prevalence was higher than the point estimate of 2.9% seroprevalence of IgM and/or IgG found in the Portuguese serological survey (ISNCOVID-19) conducted approximately in the same timeframe, but it is within its 95% CI (2.0-4.2%). It was similar to the prevalence found among those employed (3.8%; 95% CI 2.2-6.3%).[6] However, as workers were self-selected, comparisons and inferences must be cautious.

As expected, those reporting known indicators of a higher probability of being infected – known contacts with confirmed cases, ever quarantined, who had symptoms – had higher seroprevalence overall. Those with a previous negative molecular test had a higher seroprevalence than those never tested, which shows that false-negative results were likely. Also, interesting was the fact that seroprevalence increased with self-perception of increased probability of having been infected among those without a diagnosis, showing an appropriate self-assessment of risk.

One important finding is that the seroprevalence was approximately eight-times greater than reported SARS-CoV-2 infection by a molecular test, or five-times greater if we consider the true prevalence estimate. Even with considering that we may be overestimating the seroprevalence,[7] it is reasonable to expect that the SARS-CoV-2 infection was considerably more frequent than based on notified cases, particularly because testing was restricted during the initial phase of the epidemic.

No workers with an isolated IgM reactive result had a positive RT-PCR, supporting the evidence that when antibodies start being detectable the virus detection by RT-PCR is lower, and that antibody tests are appropriate to identify those previously infected but not to detect active infections,[2] although we cannot rule out the hypothesis of some false-positive IgM results.

As we had to use two different tests, though from the same manufacturer and with similar performance characteristics, error in the prevalence estimate could have occurred.Selection bias limited our ability to infer to the source population, and memory bias may have led to underreporting of exposures particularly regarding symptoms. These limitations do not seem to change the meaning of our main findings of a low seroprevalence of SARS-CoV-2 at the time of resuming working activities after the first wave of the SARS-CoV-2 infection; and that the estimated true seroprevalence was approximately five times greater than the reported SARS-CoV-2 infection burden using a molecular test information.

## Data Availability

Data may be obtained from a third party upon reasonable request sent to.

## Acknowledgements

We wish to acknowledge the team of researchers in the field. IT support from Paulo Oliveira. The health professionals from the Occupational Service, Infectious Diseases Service, and the Clinical Pathology Service from University Hospital Center São João, and from the LaBMI - Laboratory of Medical Biotechnology and Industry. We also wish to acknowledge the support from ESEP President Luís Carvalho, the P.Porto Vice-President José Barros Oliveira, and the U.Porto Vice-Rector Pedro Rodrigues.

## Data availability statement

Data may be obtained from a third party upon reasonable request sent to.

